# cGAS Expression is Enhanced in Systemic Sclerosis Associated Interstitial Lung Disease and Stimulates Inflammatory Myofibroblast Activation

**DOI:** 10.1101/2024.08.07.24311631

**Authors:** Sheeline Yu, Buqu Hu, Ying Sun, Xue Yan Peng, Chris J. Lee, Samuel Woo, John McGovern, Jana Zielonka, Tina Saber, Alexander Ghincea, Shifa Gandhi, Anjali Walia, Taylor Pivarnik, Genta Ishikawa, Shao Shuai, Huanxing Sun, Baran Ilayda Gunes, Sophia Kujawski, Stephanie Perez, William Odell, Monique Hinchcliff, John Varga, Carol Feghali- Bostwick, Maor Sauler, Jose L. Gomez, Changwan Ryu, Erica L. Herzog

**Affiliations:** Yale School of Medicine, Section of Pulmonary, Critical Care, and Sleep Medicine; Yale School of Medicine, Department of Internal Medicine, Section of Rheumatology, Allergy, and Immunology; University of Michigan, Department of Internal Medicine, Division of Rheumatology; Medical University of South Carolina, Department of Medicine; Yale School of Medicine, Department of Pathology

**Author notes:** Equally contributed to this work (Changwan Ryu, Erica L. Herzog). Corresponding author: Erica L. Herzog.

**Keywords:** Systemic Sclerosis, Interstitial Lung Disease, cGAS, inflammatory myofibroblast

## Abstract

**Objective:** The lungs of patients with Systemic Sclerosis Associated Interstitial Lung Disease (SSc-ILD) contain inflammatory myofibroblasts arising in association with fibrotic stimuli and perturbed innate immunity. The innate immune DNA binding receptor Cyclic GMP-AMP synthase (cGAS) is implicated in inflammation and fibrosis, but its involvement in SSc-ILD remains unknown. We examined cGAS expression, activity, and therapeutic potential in SSc-ILD using cultured fibroblasts, precision cut lung slices (PCLS), and a well-accepted animal model.

**Methods:** Expression and localization of cGAS, cytokines, and type 1 interferons were evaluated in SSc-ILD lung tissues, bronchoalveolar lavage (BAL), and isolated lung fibroblasts. *CGAS* activation was assessed in a publicly available SSc-ILD single cell RNA sequencing dataset. Production of cytokines, type 1 interferons, and αSMA elicited by TGFβ1 or local substrate stiffness were measured in normal human lung fibroblasts (NHLFs) via qRT-PCR, ELISA, and immunofluorescence. Small molecule cGAS inhibition was tested in cultured fibroblasts, human PCLS, and the bleomycin pulmonary fibrosis model.

**Results:** SSc-ILD lung tissue and BAL are enriched for cGAS, cytokines, and type 1 interferons. The cGAS pathway shows constitutive activation in SSc-ILD fibroblasts and is inducible in NHLFs by TGFβ1 or mechanical stimuli. In these settings, and in human PCLS, cGAS expression is paralleled by the production of cytokines, type 1 interferons, and αSMA that are mitigated by a small molecule cGAS inhibitor. These findings are recapitulated in the bleomycin mouse model.

**Conclusion:** cGAS signaling contributes to pathogenic inflammatory myofibroblast phenotypes in SSc-ILD. Inhibiting cGAS or its downstream effectors represents a novel therapeutic approach.

## INTRODUCTION

Systemic Sclerosis (SSc) is an autoimmune condition characterized by progressive fibrosis of the skin and visceral organs [1]. Patients with SSc frequently develop interstitial lung disease (SSc-ILD), a pulmonary condition characterized by inflammatory cell infiltration and fibrotic remodeling [2]. SSc-ILD underlies the considerable morbidity and mortality in patients with SSc [3], and current consensus treatment guidelines recommend immunosuppressive agents and anti-fibrotic drugs that are not universally effective and impose significant burden in terms of side effects and cost [4–6]. Thus, understanding the mechanisms contributing to both inflammation and fibrosis is imperative for the development of effective treatments to improve patient care.

The lungs of patients with SSc-ILD exhibit varying degrees of immune activation and extracellular matrix (ECM) accumulation [2]. Additionally, the lungs contain heterogeneous fibroblast populations that produce inflammatory mediators, secrete collagens and other ECM molecules, and express myofibroblast markers such as alpha smooth muscle actin (αSMA) [7–9]. These properties are inducible in normal human lung fibroblasts (NHLFs) by fibrotic stimuli, including transforming growth factor beta 1 (TGFβ1) [10] and interactions with stiffened tissue substrates [11]. The presence of inflammatory myofibroblasts in SSc-ILD may account for heterogeneous treatment responses as currently used first-line treatments are designed to either suppress the inflammatory function of immune cells or the fibrogenic function of fibroblasts. Missing from our therapeutic strategies are agents that can simultaneously mitigate inflammation and fibrogenesis in a single population of fibroblast effector cells. In view of these limitations, the identification of druggable pathways that simultaneously contribute to inflammatory responses and fibrotic functions in fibroblasts remains a critical unmet need.

Innate immune drivers of fibroblast activation have emerged as important pathogenic pathways in SSc-ILD. Of particular interest are nucleic acid receptors that recognize and respond to DNA and RNA derived from invading pathogens and injured cells. We and others have demonstrated vital roles for Toll-like receptor 9 in fibroblast activation and disease progression in SSc-ILD and associated conditions [11–13]. In contrast, the pathogenic contribution of alternate nucleic acid receptors, such as cyclic guanosine monophosphate-adenosine monophosphate synthase (cGAS), remains less well understood. The broadly expressed intracellular receptor cGAS is a nucleotidyltransferase that, upon binding to cytosolic DNA derived from invading pathogens or endogenous sources (nuclei, mitochondria), catalyzes the production of 2,3-cGAMP [14]. This substrate then activates the cytosolic kinase stimulator of interferon genes (STING) to initiate production of interferon regulatory factor-3 (IRF3) and nuclear factor-kB (NFkB) dependent proinflammatory cytokines, chemokines, and type I interferons in immune cells [15]. Reflecting the emerging importance of cGAS in disease development, numerous approaches to cGAS-STING modulation exist in the therapeutic pipeline for conditions of chronic inflammation and autoimmunity [15, 16]. Intriguingly, the cGAS-STING pathway has also been implicated as a fibroblast activator in diverse conditions such as cancer [17], aging [18], pulmonary fibrosis [19], and SSc [20]; and endogenous cGAS ligands such as DNA from mitochondria (mtDNA) and nuclear sources are elevated in patients with SSc [12, 13, 20]. However, the mechanistic and therapeutic potential of cGAS in SSc-ILD is currently unknown.

On the basis of this biology, we surmised that engagement of cGAS drives inflammatory myofibroblast accumulation in SSc-ILD. To test this hypothesis, we evaluated the expression and function of cGAS in patient samples, *ex vivo* modeling systems, and in a well-accepted mouse model. Our results show that expression of cGAS and its associated inflammatory mediators is increased in lung tissue and bronchoalveolar lavage (BAL) fluid from SSc-ILD patients, where it is constitutively expressed and activated in SSc-ILD fibroblasts, and inducible in NHLFs following stimulation with either TGFβ1 or mechanical stimuli. In these settings, and in human precision cut lung slices (PCLS), cGAS expression is paralleled by production of inflammatory mediators and αSMA expression that are mitigated by a small molecule cGAS inhibitor. Similar anti-fibrotic effects of cGAS inhibition are seen in an experimental model of mouse lung fibrosis. Altogether, our compelling results implicate cGAS in inflammatory fibroblast activation and fibrosis underlying SSc-ILD pathogenesis, putting forth cGAS inhibition as a novel therapeutic in this disease.

## MATERIALS AND METHODS

Detailed methods are provided in the online supplement.

### Human Subjects

De-identified lung tissues and primary lung fibroblasts that had been previously obtained at the University of Pittsburgh (IRB 970946), and de-identified BAL fluid specimens that had been previously collected from healthy donors and patients with SSc using informed consent on protocols approved by Institutional Review Board at the Yale School of Medicine (HIC 2000024862), were used in this study.

### Single Cell RNA Sequencing (scRNA seq) Analysis

Analyses of scRNA seq data (GSE128169) were performed with Seurat version 5.0.318 as described in the online supplement. To identify cell types expressing genes associated with cGAS, the AddModuleScore, a Seurat function that calculates the gene expression level of a cluster of genes on a single cell level and then subtracts the aggregated expression of a randomly assigned control set, was used.

### Fibroblast Cultures

Fibroblasts harvested from the lungs of patients with SSc-ILD were cultured in the absence or presence of the small molecule cGAS inhibitor G140 (10 µM, Invivogen). NHLFs were cultured in the absence or presence of recombinant human TGFβ1 (5 ng/ml) and CpG ODN2006 (50 µmol) and in the absence or presence of G140. In parallel, NHLFs were cultured on collagen-coated hydrogels (Matrigen) of 1 kPA and 25 kPA stiffness in the absence or presence of G140.

### Human PCLS

PCLS from normal human donors (Institute for In Vitro Sciences) were incubated in culture media supplemented with or without an inflammatory fibrotic cocktail (IFC) comprised of the following: TGFβ1 (5 ng/ml), platelet-derived growth factor-AB (PDGF-AB, 5 μM), tumor necrosis factor alpha (TNF-α, 10 ng/ml), lysophosphatidic acid (LPA, 5 μM), and CpG ODN2006 (50 µmol); PCLS subjected to this cocktail were then treated in the absence or presence of G140 (10 nM).

### Mouse Experiments

Animal procedures were approved by the Yale Institutional Animal Care and Use Committee. C57Bl/6 mice received a single dose of inhaled bleomycin (0.8 units/kg) or normal saline. Vehicle control or G140 (30 mg/kg) was administered via intraperitoneal injection on days 3, 7, and 10 post-bleomycin. Mice were euthanized on day 14 and biospecimens were collected for analysis.

### RNA Isolation and qRT-PCR

Total cellular RNA was isolated and reverse transcribed for *GAPDH*, *ACTA2,* and *CGAS* from biospecimens as described in the online supplement.

### Cell Lysis

Cells were washed and lysed in a solution containing Pierce RIPA buffer and a Halt™ Protease Inhibitor Cocktail (ThermoFisher).

### Quantification of Cytokines

Multiplex quantification of interleukin-6 (IL-6), interleukin 1-beta (IL-1β), tumor necrosis factor alpha (TNF-α), monocyte chemoattractant protein-1 (MCP-1), interferon-gamma inducible protein-10 (IP-10), interferon alpha (IFN-α), and interferon beta (IFN-β) using the U-PLEX Cytokine Panel Human Kit or U-Plex Cytokine Panel Mouse Kit (Mesoscale Discovery).

## ELISA

Cell lysates were assayed for αSMA (Abcam) or cGAS (Cell Signaling Technology) via commercially available ELISA as described in the online supplement.

### MTT Assay

Cell viability was determined using MTT assays (Thermofisher) as described in the online supplement.

### cGAS Immunohistochemistry (IHC)

Formalin fixed, paraffin embedded slides of human lung tissue from healthy or SSc-ILD donors (n=4/each) were stained with a human cGAS antibody (1:100, Invitrogen) as described in the online supplement. cGAS quantification was performed by manually counting total nuclei and nuclei from cGAS-expressing cells; results were reported as the percentage of positive cGAS cells per high-power field.

### Immunofluorescence (IF)

IF detection of αSMA and cGAS in fibroblasts were performed with mouse anti-αSMA (1:250, Abcam) and rabbit monoclonal cGAS (1:100, Invitrogen) antibodies as described in the online supplement.

### Lung Histology

Formalin fixed, paraffin embedded lung sections from PCLS or mouse lungs were stained with Masson’s trichrome [21]. Collagen was quantified with ImageJ software as described in the online supplement. Soluble collagen was quantified using the Sircol Collagen Assay kit (Biocolor) [22].

### Bulk RNA Sequencing

RNA isolated from PCLS was sequenced on the Illumina NovaSeq platform as described in the online supplement. Differential gene expression was identified with the DESeq2 package on R statistical software (version 4.3.2). Heatmaps were generated in R with the gplots package. Pathway enrichment analysis was conducted via MetaCore; a false discovery rate (FDR) < 0.10 was considered significant.

### Statistical Analysis

Data distribution was assessed using Shapiro-Wilk test, and categorical data was analyzed with Fisher’s exact test using GraphPad Prism 9.4.1. Multiple hypothesis testing was conducted with one-way ANOVA with Bonferroni correction with GraphPad Prism. Pairwise comparisons were conducted with an unpaired t-test or Mann Whitney test based on data distribution with GraphPad Prism. p-values < 0.05 were considered significant.

## RESULTS

### Expression of cGAS and its associated soluble mediators are enriched in the lungs of patients with SSc-ILD

To assess a potential relationship between cGAS and SSc-ILD, IHC was performed on lung tissues obtained from control (unsuitable organ donors) or SSc-ILD patients undergoing orthotopic lung transplantation. In contrast to the low level of cGAS detection observed in control samples, SSc-ILD lung tissues displayed abundant cGAS expression in cells exhibiting the morphology of immune populations, epithelium, and fibroblasts (Figures 1A,B). cGAS-associated soluble mediators were then measured in BAL specimens from control and SSc donors shown in Table 1. Compared to healthy controls, SSc-ILD BAL showed high concentrations of cytokines (IL-6, IL-1β, TNF-α, Figures 1C-E), chemokines (MCP-1, IP-10, Figures 1F,G), and type 1 interferons (IFN-α, IFN-β, Figures 1H,I) all of which are known to be downstream of cGAS activation. These data show that the lungs of patients with SSc-ILD show strong expression of cGAS and its associated soluble mediators.

**Figure 1.**
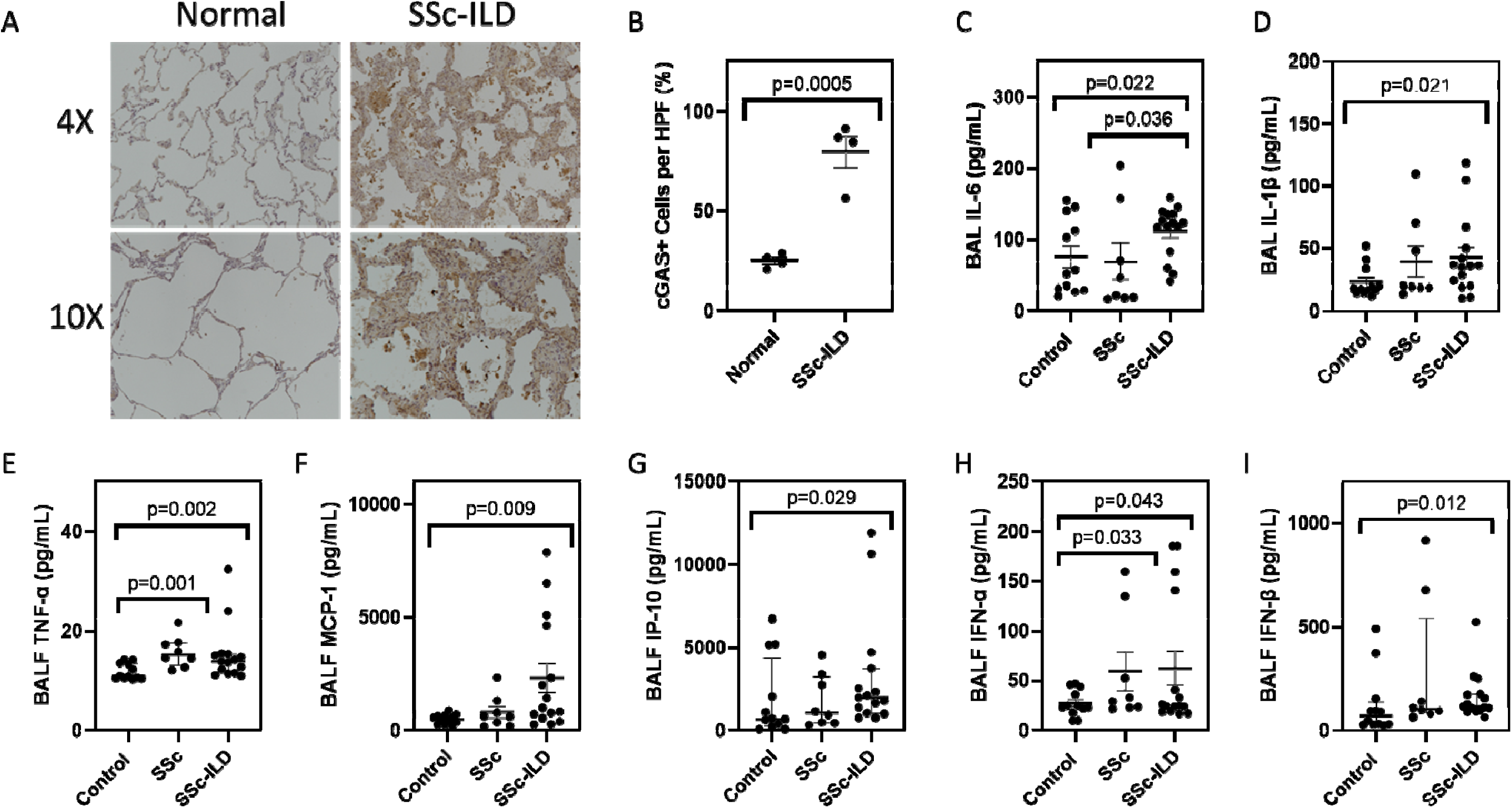
SSc-ILD lungs show enhanced expression of cGAS and its associated inflammatory mediators. (**A**) IHC for cGAS showed enhanced detection in the SSc-ILD lung (right) as compared to the normal lung (left). **(B)** A higher percentage of cGAS positive cells (per high-powered field) was observed in the SSc-ILD lung. BAL fluid samples collected from healthy donors and SSc patients showed that relative to healthy participants, SSc-ILD patients exhibited increased concentrations of **(C)** IL-6, **(D)** IL-1β, **(E)** TNF-α, **(F)** MCP-1, **(G)** IP-10, **(H)** IFN-α, and **(I)** IFN-β.

**Table 1.** Demographic and clinical characteristics of study participants.

|  | Healthy controls | SSc without ILD | SSc-ILD |
| --- | --- | --- | --- |
| N | 12 | 8 | 15 |
| Age (mean ± SD) | 53.04 ± 14.97 | 51.89 ± 10.67 | 60.10 ± 9.70 |
| Gender (n, % female) | 6 (50.0%) | 8 (100%) | 10 (66.7%) |
| Race |  |  |  |
| White (n, %) | 9 (75.0%) | 6 (75%) | 12 (80.0%) |
| Black (n, %) | 3 (25.0%) | 1 (12.5%) | 2 (13.3%) |
| Other (n, %) | 0 (0%) | 1 (12.5%) | 1 (6.7%) |
| Smoking |  |  |  |
| Ever (n, %) | 2 (16.7%) | 3 (37.5%) | 7 (44.7%) |
| Never (n, %) | 10 (83.3%) | 5 (62.5%) | 8 (53.3%) |
| SSc Subtype |  |  |  |
| Diffuse (n, %) | N/A | 1 (12.5%) | 5 (33.3%) |
| Limited (n, %) | N/A | 7 (87.5%) | 10 (66.7%) |
| Disease Duration (mean ± SD) | N/A | 4.50 ± 2.77 | 6.44 ± 3.71 |
| MRSS (median ± IQR) | N/A | 4.5 ± 6.75 | 7.0 ± 3.00 |
| Scl-70 (n, %) | N/A | 2 (25%) | 7 (46.7%) |
| Centromere (n, %) | N/A | 5 (62.5%) | 3 (20.0%) |
| RNA Pol III (n, %) | N/A | 0 (0%) | 3 (20.0%) |
| None (n, %) | N/A | 1 (12.5%) | 2 (13.3%) |
| FVC% (mean ± SD) | N/A | 90.00 ± 18.98 | 80.22 ± 19.19 |
| DLCO% (mean ± SD) | N/A | 91.14 ± 22.83 | 64.89 ± 24.80 |

### Enhanced expression and activation of cGAS in SSc-ILD fibroblasts

Having identified enhanced cGAS expression in SSc-ILD lung tissues, we then investigated this biology in disease-relevant cell types. We began by analyzing a publicly available scRNA seq dataset (GSE128169) generated from end-stage SSc-ILD lung explants. Consistent with our IHC findings above, multiple cell populations showed transcriptional evidence of cGAS activation (Figures S1 and 2A). When this analysis was limited to stromal cells, fibroblasts exhibited high expression of cGAS pathway transcripts (Figure 2B). These *in silico* studies were confirmed *in vitro* with primary lung fibroblasts, where evaluation of cGAS expression via qRT-PCR and ELISA showed increased expression at the mRNA and protein levels (Figures 2C,D). Moreover, IF analysis revealed that the baseline nuclear cGAS localization seen in NHLFs was accompanied by the detection of cytoplasmic aggregates in SSc-ILD fibroblasts, indicating that these cells may exist in a state of heightened cGAS activation (Figures S2 and 2E). Importantly, this phenotype was accompanied by increased production of cGAS-associated inflammatory mediators (IL-6, IL-1β, TNF-α, MCP-1, IP-10, IFN-α, IFN-β) along with elevated mRNA and protein levels of the myofibroblast marker αSMA (Figures 3A-I). These results implicate cGAS as a driver of inflammatory myofibroblast activation in SSc-ILD.

**Figure 2.**
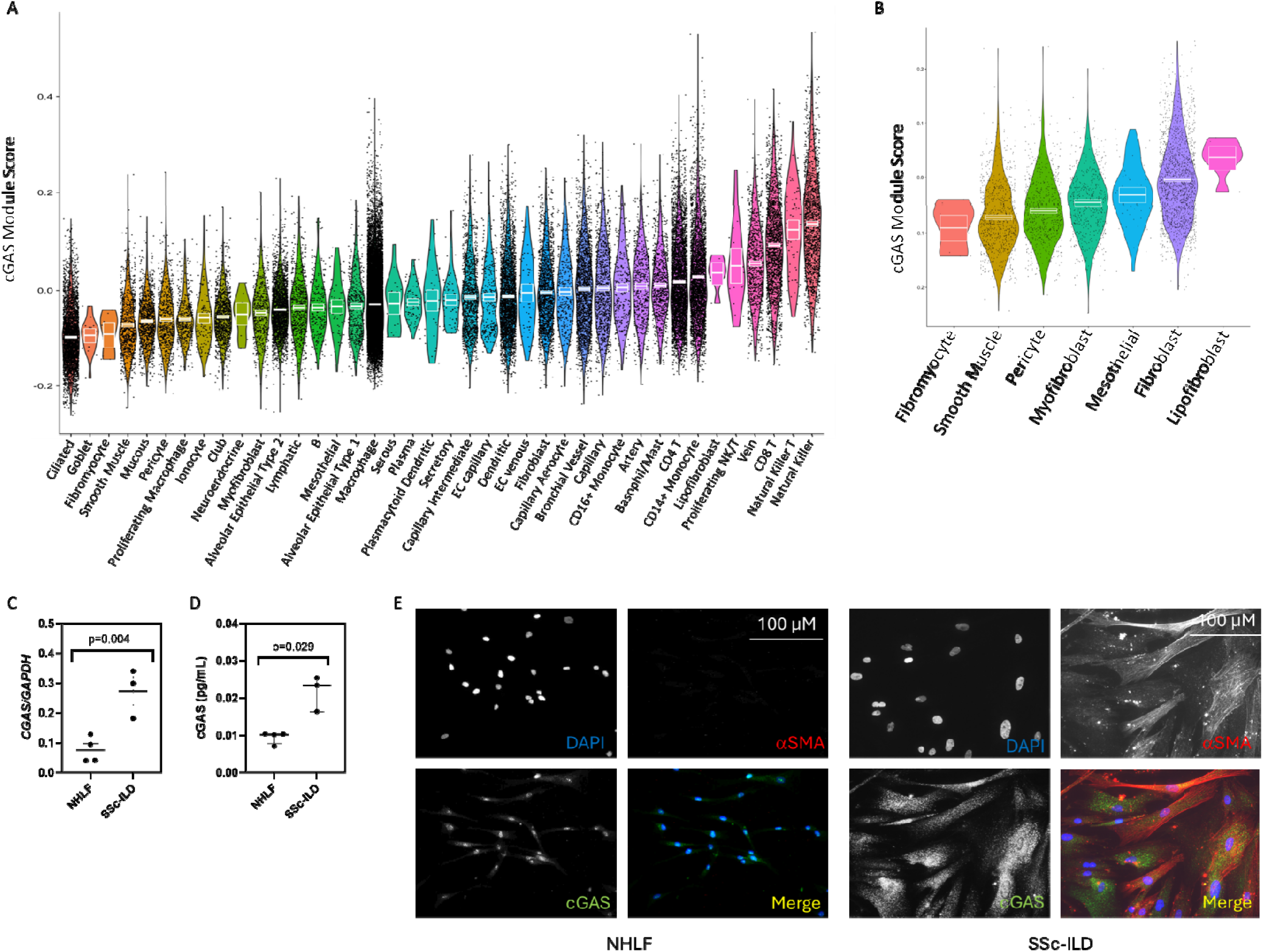
SSc-ILD lung fibroblasts are enriched for cGAS expression. (**A**) Violin plot of single cell RNA sequencing analysis (derived from GEO dataset GSE128169) from the lungs of patients with SSc-ILD demonstrating median expression of cGAS-related genes across multiple cell populations, which was highest among immune and endothelial cells. **(B)** Among stromal cells, fibroblasts (purple) demonstrated enhanced expression of these genes. **(C)** qRT-PCR and **(D)** ELISA measurements of NHLFs and SSc-ILD fibroblasts showed increased transcription and protein expression of cGAS in SSc-ILD fibroblasts. **(E)** IF evaluation of NHLFs (left) and SSc-ILD fibroblasts (right) showed the increased presence of αSMA and cytoplasmic aggregation of cGAS in SSc-ILD fibroblasts (versus the nuclear localization of cGAS in NHLFs). Scale bar = 100 microns.

**Figure 3.**
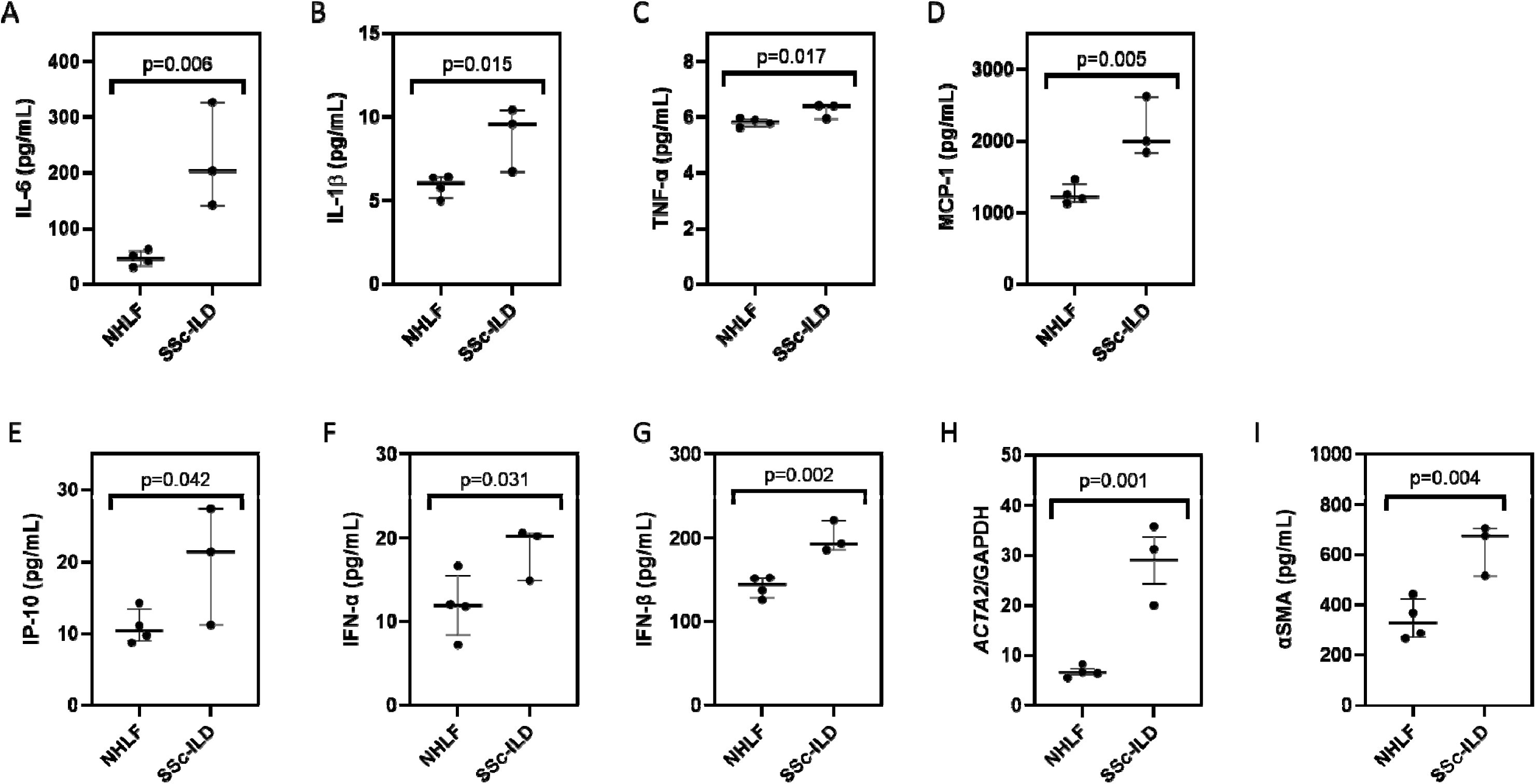
SSc-ILD fibroblasts express high concentrations of cGAS associated soluble mediators and αSMA. As compared to NHLFs, SSc-ILD fibroblasts expressed high protein levels of **(A)** IL-6, **(B)** IL-1β, **(C)** TNF-α, **(D)** MCP-1, **(E)** IP-10, **(F)** IFN-α, and **(G)** IFN-β. Relative to normal cells, SSc-ILD fibroblasts expressed increased αSMA **(H)** transcripts via RT-qPCR and **(I)** protein via ELISA.

### A small molecule cGAS inhibitor mitigates inflammatory mediator production and fibrotic activation of SSc-ILD fibroblasts

To link cGAS more firmly with inflammatory myofibroblast activation in SSc-ILD, we performed loss of function studies using the cGAS inhibitor G140. This commercially available, highly specific small molecule inhibitor blocks DNA entry into human cGAS catalytic pocket to impede enzymatic activation [23]. In our study, treatment of SSc-ILD fibroblasts with G140 reduced cytoplasmic cGAS aggregates (Figure 4A), suggesting successful inhibition. This observation was paralleled by decreased production of inflammatory mediators (IL-6, IL-1β, TNF-α, MCP-1, IP-10, IFN-α, and IFN-β, Figures 4B-H). G140 treatment also reduced αSMA mRNA measured by qRT-PCR (Figure 4I) and protein measured by IF and ELISA (Figures 4A,J). Importantly, these findings were not due to alterations in cellular viability as the results of an MTT assay were similar between untreated and treated cells (Figure 4K). These studies show that cGAS inhibition reduces production of cytokines, chemokines, type 1 interferons, and αSMA expression in SSc-ILD fibroblasts, providing proof of concept evidence for targeting cGAS in this disease.

**Figure 4.**
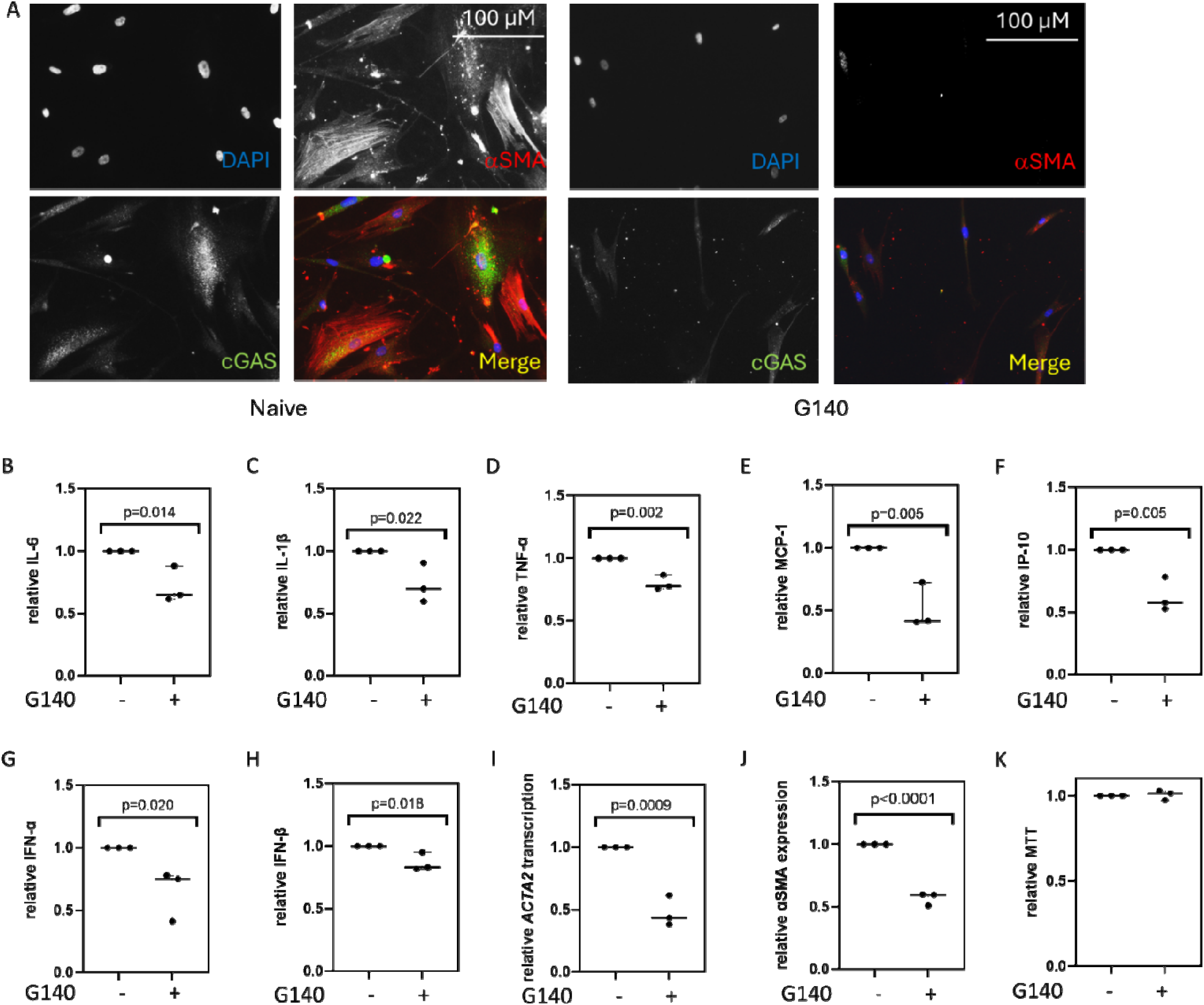
Small molecule cGAS inhibition mitigates inflammatory fibrotic respon es in SSc-ILD fibroblasts. (**A**) IF staining of naïve (left) and G140 treated (right) SSc-ILD fibroblasts showed reduced detection of both cytoplasm c cGAS aggregates and αSMA in treated cells. Scale bar = 100 microns. These G140-treated cells displayed decreased expression of **(B)** IL-6, **(C)** IL-1β, **(D)** TNF-α, **(E)** MCP-1, **(F)** IP-10, **(G)** IFN-α, and **(H)** IFN-β. Treated cells also displayed a reduction in the **(I)** transcription and **(J)** protein expression of αSMA. **(K)** An MTT assay showed that G140 did not influence the viability of these fibroblasts. Data was normalized to the untreated condition.

### Small molecule cGAS inhibition suppresses TGF**β**1 and CpG induced fibroblast activation

Studies of diseased lung fibroblasts are useful for the development of mechanistic-based SSc-ILD therapies. However, while providing evidence of target engagement, they are limited by the end-stage nature of the disease. Because the therapeutic goal is to prevent clinical progression, studies aimed at preventing cellular activation have the potential to halt or even reverse disease. Reasoning that fibroblasts adopt their ultimate phenotype in response to microenvironmental cues, we developed a model of cGAS activation in which NHLFs were co-stimulated with TGFβ1 and CpG ODN2006 to simulate the endogenous cGAS ligand mtDNA (Figures S3A,B). Following co-stimulation, NHLFs phenocopied many aspects of SSc-ILD fibroblasts, including increased expression and cytoplasmic aggregation of cGAS (Figures S4A-C), production of inflammatory mediators (IL-6, IL-1β, TNF-α, MCP-1, IP-10, IFN-α, IFN-β, Figures S4D-J), and αSMA mRNA and protein (Figures S4K,L). These changes were cGAS-dependent as treatment with G140 diminished cytoplasmic cGAS aggregation (Figure 5A) and suppressed production of all inflammatory mediators except for IL-6 (Figures 5B-H). Production of αSMA mRNA and protein were also reduced (Figures 5I,J). Lastly, a MTT assay revealed these findings to be unrelated to changes in cell viability (Figure 5K). Taken together, these data provide evidence of cGAS antagonism as a novel strategy of mitigating the fibroblast activation that arises in response to TGFβ1 and endogenous cGAS ligands.

**Figure 5.**
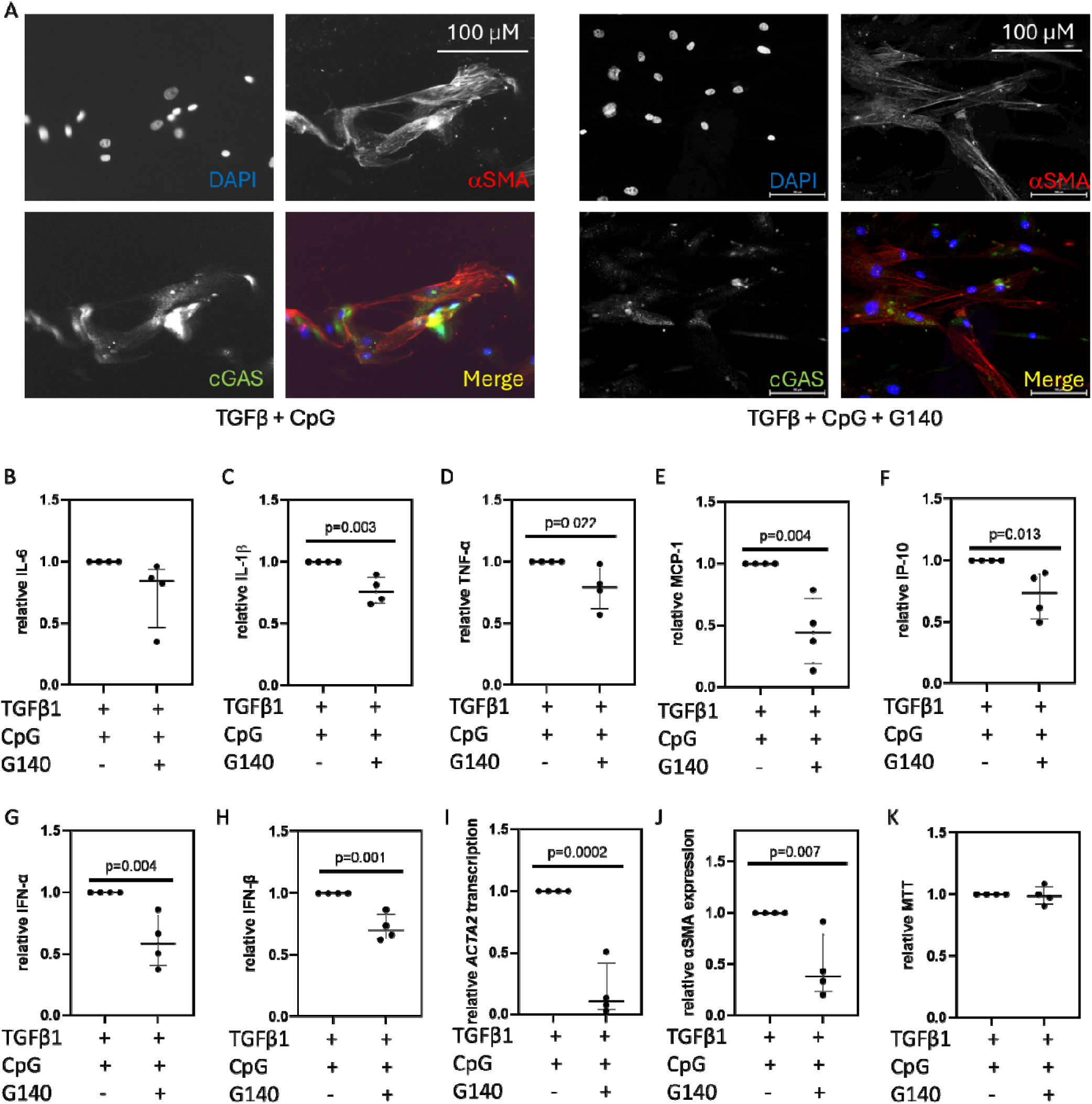
A small molecule cGAS inhibitor suppresses inflammatory fibroblast activation induced by TGFβ1 and CpG co-stimulation. (**A**) IF of NHLFs co-stimulated with TGFβ1 and CpG in the absence (left) or presence (right) of G140 showed that treatment diminished the presence of cGAS aggregates in the cytoplasm. Scale bar = 100 microns. Although treatment with G140 did not change levels of **(B)** IL-6, it attenuated expression of **(C)** IL-1β, **(D)** TNF-α, **(E)** MCP-1, **(F)** IP-10, **(G)** IFN-α, and **(H)** IFN-β, as well as **(I)** mRNA transcription and **(J)** protein expression of αSMA. **(K)** An MTT assay showed that these findings were unrelated to changes in cell viability. Data were normalized to the TGFβ1 + CpG condition.

### A small molecule cGAS inhibitor opposes stiffness-induced fibroblast activation

Cells encounter and respond to physical properties, such as stiffness, in their microenvironment. While cGAS has been previously linked to fibrotic mechanosensing [18], its involvement in SSc-ILD and therapeutic targeting in lung fibroblasts remains underexplored. Thus, we investigated this area using tunable hydrogels constructed to simulate the stiffness of normal (1 kPA) or SSc-ILD (25 kPA) lung [24]. Compared to cells cultured under physiologic conditions, NHLFs grown on 25 kPA gels developed increased cGAS mRNA transcription and protein levels (Figures S5A,B), secretion of IL-1β, MCP-1, IP-10, IFN-α, and IFN-β, but not IL-6 or TNF-α (Figures S5C-I) as well as induction of αSMA mRNA and protein expression (Figures S5J,K). Treatment with G140 reduced IL-1β, IP-10, IFN-α, and IFN-β, but not MCP-1 (Figures 6A-E), and attenuated αSMA mRNA and protein (Figures 6F,G) without affecting cellular viability (Figure 6H). Overall, these data frame cGAS as a druggable driver of mechanosensitive fibroblast responses that are pathogenic in SSc-ILD.

**Figure 6.**
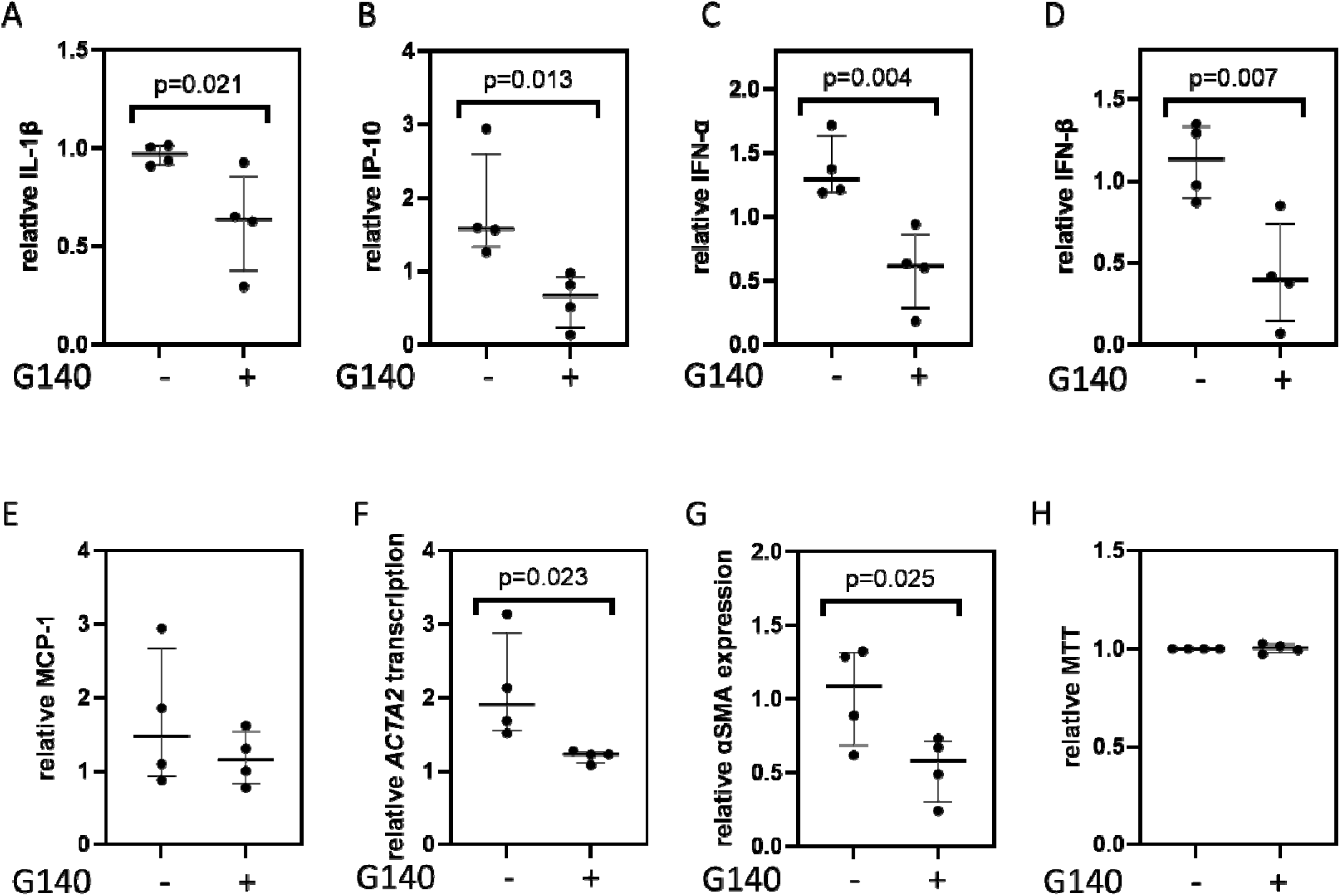
A small molecule cGAS inhibitor mitigates stiffness-induced fibroblast activation. NHLFs were cultured on tunable hydrogels that approximate the stiffness of the normal (1kPA) and fibrotic (25 kPA) lung. As compared to NHLFs cultured in the untreated 25 kPA condition, those grown on 25 kPA hydrogels treated with G140 exhibited decreased levels of **(A)** IL-1β, **(B)** IP-10, **(C)** IFN-α, and **(D)** IFN-β; expression of **(G)** MCP-1 remained unchanged. This inhibitor also led to a reduction in the **(F)** transcription and **(G)** expression of αSMA. **(H)** An MTT assay showed that these findings were unrelated to changes in cell viability. Data was normalized to their respective untreated or treated 1 kPA condition.

### A small molecule cGAS inhibitor suppresses inflammatory mediator production and fibrotic endpoints in human PCLS

Studies of isolated lung fibroblasts provide direct evidence of cell autonomous responses in these fibrotic cells. However, they are limited by the lack of information regarding the complex multicellularity of the adult lung, which can activate fibroblasts via non-cell autonomous processes. To overcome these issues, we extended our studies to include PCLS, an advanced *in vitro* modeling system that allows previously impossible tissue level insights into human lung injury, repair, and remodeling. This technology improves upon standard primary monoculture methods by replicating the complex 3D topography and multicellularity of the lung [25]. Our preliminary studies used a model of normal PCLS cultured in media containing an inflammatory fibrotic cocktail (IFC) of TGFβ, PDGF-AB, TNF-α, and LPA, and supplemented with the mtDNA surrogate CpG ODN2006. As compared to naïve PCLS, those cultured in this IFC displayed enhanced collagen content (Figures S6A-B), cGAS transcripts (Figure S6C), protein levels of IL-6, IL-1β, TNF-α, MCP-1, IP-10, and IFN-α, but not IFN-β (Figures S6D-J). αSMA transcripts were also increased (Figure S5K). Treatment with the cGAS inhibitor G140 reduced all these endpoints (Figures 7A-I). Lastly, to identify parallel transcriptional changes that occur in the absence or presence of G140, we conducted exploratory bulk RNA sequencing and identified 126 differentially expressed genes (Figure 7J). Unbiased pathway enrichment analysis revealed a multitude of transcriptional changes involving inflammatory and immune function with the most highly altered pathways being related to cytoskeletal remodeling, IL-17, cell adhesion, antigen presentation, NF-kB activation, interferon beta signaling, and mesenchymal cell activation (Figure 7K). The finding that cGAS inhibition suppresses production of cytokines, chemokines, type 1 interferons, αSMA expression, and collagen production during human lung fibrogenesis suggests that targeting cGAS and/or its associated pathways holds therapeutic potential in SSc-ILD.

**Figure 7.**
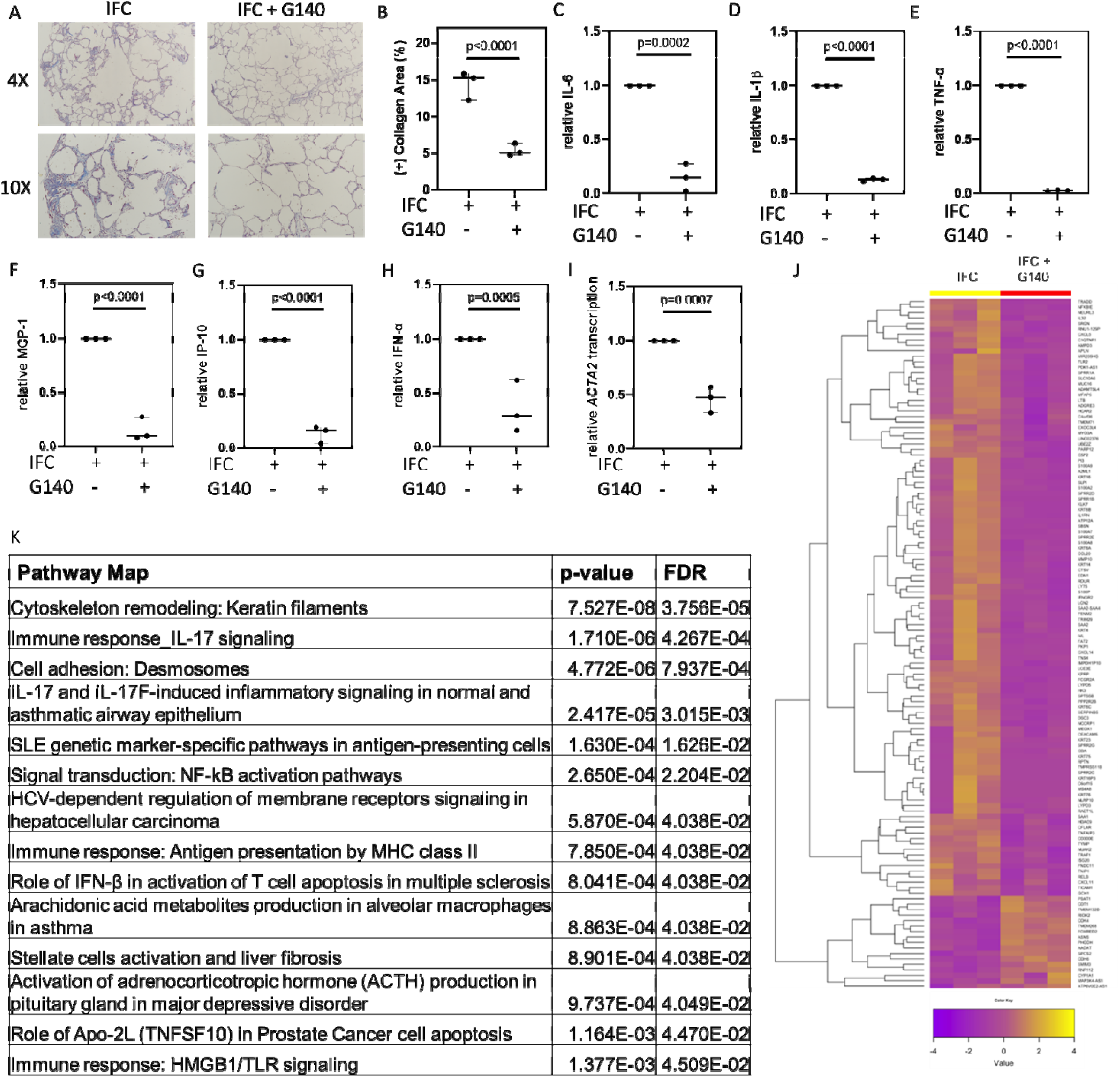
A small molecule cGAS inhibitor abrogates inflammatory fibrotic responses in human precision cut lung slices. (**A**) Representative images of Masson’s Trichrome staining of human PCLS sections from those subjected to an inflammatory fibrotic cocktail (IFC) in the absence (left) or presence (right) of G140. **(B)** Collagen quantification via ImageJ software revealed a reduction in collagen content following treatment with G140. Treatment with this inhibitor mitigated expression of **(C)** IL-6, **(D)** IL-1β, **(E)** TNF-α, **(F)** MCP-1, **(G)** IP-10, and **(H)** IFN-α, and transcription of **(I)** *ACTA2*. Data were normalized to the IFC condition. **(J)** Heatmap of bulk RNA sequencing between IFC cultured PCLS in the absence or presence revealed 126 differentially expressed genes (DEGs). **(K)** Unbiased pathway enrichment analysis.

### A small molecule cGAS inhibitor mitigates bleomycin-induced pulmonary fibrosis

Finally, having demonstrated the benefit of a cGAS inhibitor in orthogonal *ex vivo* modeling experiments, we sought to determine its efficacy *in vivo*. For this purpose, we used the bleomycin model of fibrosis in mice, which is the most widely accepted SSc-ILD animal model [2]. Mice were subjected to orotracheal instillation of either normal saline or bleomycin in presence of vehicle or G140. As shown in Figures S7A-C, bleomycin challenged mice exhibited greater lung collagen accumulation than those that received normal saline or G140 alone. Evaluation of BAL fluid revealed elevations of IL-6, IL-1β, TNF-α, MCP-1, IP-10, IFN-α, and IFN-β (Figures S7D-J). When bleomycin-challenged mice were treated with G140, we observed marked reductions in lung collagen content (Figures 8A-C) and BAL concentrations of IL-6, TNF-α, IP-10, IFN-α, and IFN-β, but not IL-1β and MCP-1 (Figures 8D-J). When viewed in combination, our results identify cGAS as a modulator of inflammatory fibroblast activation in mammalian lung fibrosis and supports the clinical development of cGAS inhibitors for treatment of SSc-ILD.

**Figure 8.**
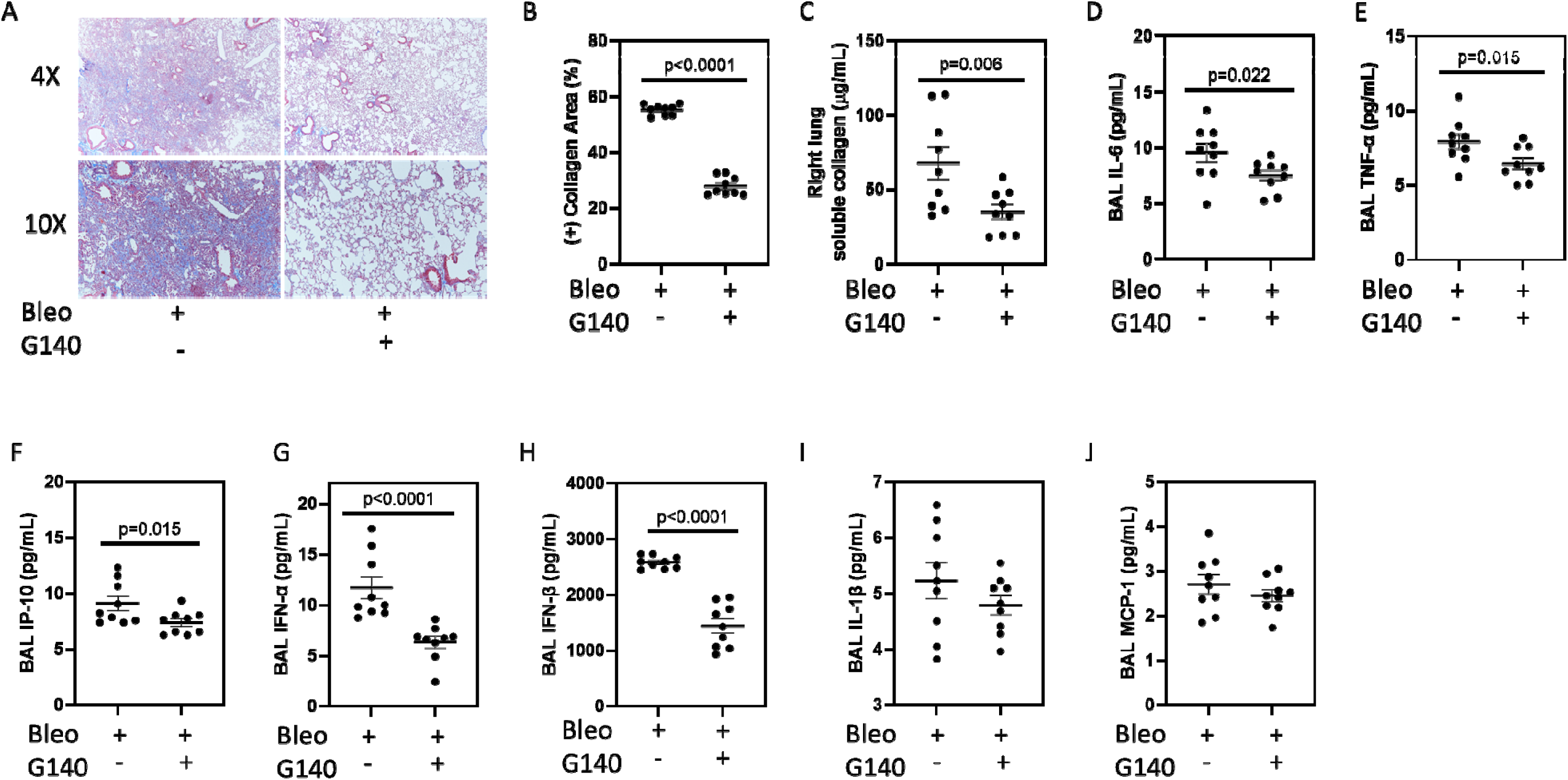
A small molecule cGAS inhibitor ameliorates bleomycin induced pulmonary fibrosis. (**A**) Representative images of Masson’s Trichrome staining of lung sections from bleomycin challenged mice in the absence (left) or presence (right) of G140. Collagen quantification via **(B)** ImageJ software and **(C)** Sircol assay revealed a reduction in collagen content following treatment with G140. Bleomycin exposed mice tre ted with G140 displayed reduced BAL concentrations of **(D)** IL-6, **(E)** TNF-α, **(F)** IP-10, **(G)** IFN-α, and **(H)** IFN-β, while levels of **(H)** IL-1β and **(J)** MCP-1 remained unchanged.

## DISCUSSION

Innate immunity and inflammatory myofibroblast activation are increasingly linked to SSc-ILD; however, their mechanistic and therapeutic implications are not well understood [26]. Here we identified the cytosolic DNA sensor cGAS as a mechanistic driver of and therapeutic target in SSc-ILD. Our results revealed that expression of cGAS and its downstream mediators are enhanced in SSc-ILD lung tissues, BAL fluid, and lesional fibroblasts. These findings are recapitulated in SSc-ILD modeling systems, including NHLFs exposed to biochemical and biophysical fibrotic stimuli, human PCLS exposed to fibrogenic mediators, and the well accepted bleomycin mouse model. A small molecule cGAS inhibitor exerted an anti-inflammatory and antifibrotic effects in all these settings. Targeting aberrant cGAS activation holds promise as an innovative strategy in SSc-ILD, as well for myriad other inflammatory fibrosing conditions characterized by autoimmunity [27], neurodegeneration [28], cancer [17], and aging [18].

Upon its 2013 discovery, cGAS was described as a nucleotidyltransferase that recognizes pathogen-derived DNA and activates type 1 interferon production in a STING-dependent, IRF3 dependent manner [14]. Since this time, cGAS has been shown to be activated by endogenous DNA, including that derived from nuclei and mitochondria [10]. cGAS can recognize self-ligands that leak into the cytoplasm following DNA damage responses or mitochondrial stress, or that which have been internalized as damage associated molecular patterns (DAMPS) present in the extracellular milieu. Our prior work implicated mtDNA as a putative cGAS activator [10] and the present study advances this notion as we were able to induce fibroblast activation and cGAS signaling with the administration of exogenous CpG, a surrogate for cell-free mtDNA. Further work characterizing the nature of mtDNA and its interactions with cGAS has the potential to elucidate intervenable innate immune mechanisms of inflammatory fibroblast activation in SSc-ILD.

The results reported herein are predicated on the ability of cGAS to activate STING and induce NFkB and IRF3 signaling [14]. In our work, cGAS inhibition suggested context dependent aspects of the various modeling systems. For example, while SSc-ILD BAL and fibroblasts are enriched for NFkB-dependent and IRF3-dependent cytokines and type 1 interferons, we have not characterized the contribution of STING and its associated transcription factors in our work. The expression of NFkB and IRF3 dependent inflammatory mediators supports pleiotropic effect, and the effect on αSMA expression likely reflects the recently described role of cGAS-STING in canonical TGFβ1 activation and YAP/TAZ mediated mechanosensing. It is equally possible, however, that our observations stem from noncanonical cGAS pathways, such as PERK, that are independent of STING. Further study is needed to decipher the context-dependent paradigm of cGAS activation and its downstream signaling pathways in SSc-ILD and related conditions.

In terms of therapeutic potential, cGAS inhibition remains an active area of investigation for clinical benefit. Currently available antagonists include the anti-malarial agents hydroxychloroquine and quinacrine, which bind to the minor groove of the DNA-cGAS interface to disrupt ligand-receptor interactions [29]. Suramin, a drug indicated for the treatment of river blindness and African sleeping sickness, acts as a DNA mimic that competitively binds to cGAS and inhibits 2,3,-cGAMP synthesis [30]. While repurposing these agents for SSc-ILD is a consideration, the potential for non-specific, off-target effects limits their widespread use [31, 32]. Thus, the identification of compounds that directly inhibit cGAS has been actively pursued. One promising compound was RU.521, which occupies the catalytic site of cGAS to inhibit generation of 2,3,-cGAMP [31]. However, this effect was only seen in mouse studies, propelling the development of G140, a human-specific small molecule inhibitor of cGAS with a mechanism of action similar to RU.521 [23]. Moreover, relative to similar compounds, G140 demonstrated superior efficacy inhibiting production of various lipopolysaccharide-induced cytokines in mouse lung injury models [32]. Our study is the first to evaluate G140 in this inflammatory fibrotic setting, and further work is needed for testing this agent in SSc-ILD human cohorts.

While novel and thought-provoking, our findings raise additional questions for future research. Although we have identified fibroblasts as one site of activated cGAS function, additional cell types are likely involved. Mechanistically, we have not determined if cGAS activation mediates canonical versus non-canonical TGFβ activation, nor its interactions with mechanosensitive signaling, such as those involving Hippo [33], YAP [34], FAK [34], and Rho associated kinase [35]. In terms of the cGAS ligand, we have not determined the contribution of DNA derived from mitochondrial, nuclear, or even microbial sources. Moreover, we have yet to determine the impact of canonical cGAS signaling components, such as the second messenger protein 2,3,-cGAMP and STING. Lastly, we have not evaluated how cGAS inhibition compares to immunosuppressive strategies [5] and anti-fibrotic agents [36] that are currently in use. Despite these relatively minor limitations, in the first of its kind study, we found that cGAS is expressed in SSc-ILD lung tissues and that its inhibition mitigates the development of inflammatory fibrosis in cellular, tissue-based, and animal models. Further investigation may yield the development of cGAS targeted therapies for SSc-ILD and related conditions.

## Data Availability

All data produced in the present study are available upon reasonable request to the authors.

## ACKNOWLEDGMENTS

To all of our patients with scleroderma: the authors commend your bravery and resilience in combatting this relentless disease. We hope and encourage you all to live your best lives.

## Contributions

SY conducted the investigation, interpreted data, and wrote and edited the manuscript. BH, YS, XYP, CJL, SW, JM, JZ, TS, AG, SG, AW, TP, SS, HS, MS, and JLG performed the investigation and analyzed data. BIG, SK, SP, WO, and MH recruited subjects, performed the investigation, and analyzed data. JV provided conceptualization, resources, supervision, and funding acquisition. CFB provided conceptualization and resources. CR and ELH provided conceptualization, resources, supervision, project administration, funding acquisition, and wrote and edited the manuscript. All authors participated in manuscript preparation and provided final approval of the submitted work.

## TAKE HOME MESSAGE

Using various translational models of SSc-ILD, we found that activation of the innate immune receptor cGAS contributes to inflammatory myofibroblast phenotypes that are attenuated by inhibiting this receptor.

