## Supplemental Figure 1 for "cGAS Expression is Enhanced in Systemic Sclerosis Associated Interstitial Lung Disease and Stimulates Inflammatory Myofibroblast Activation"

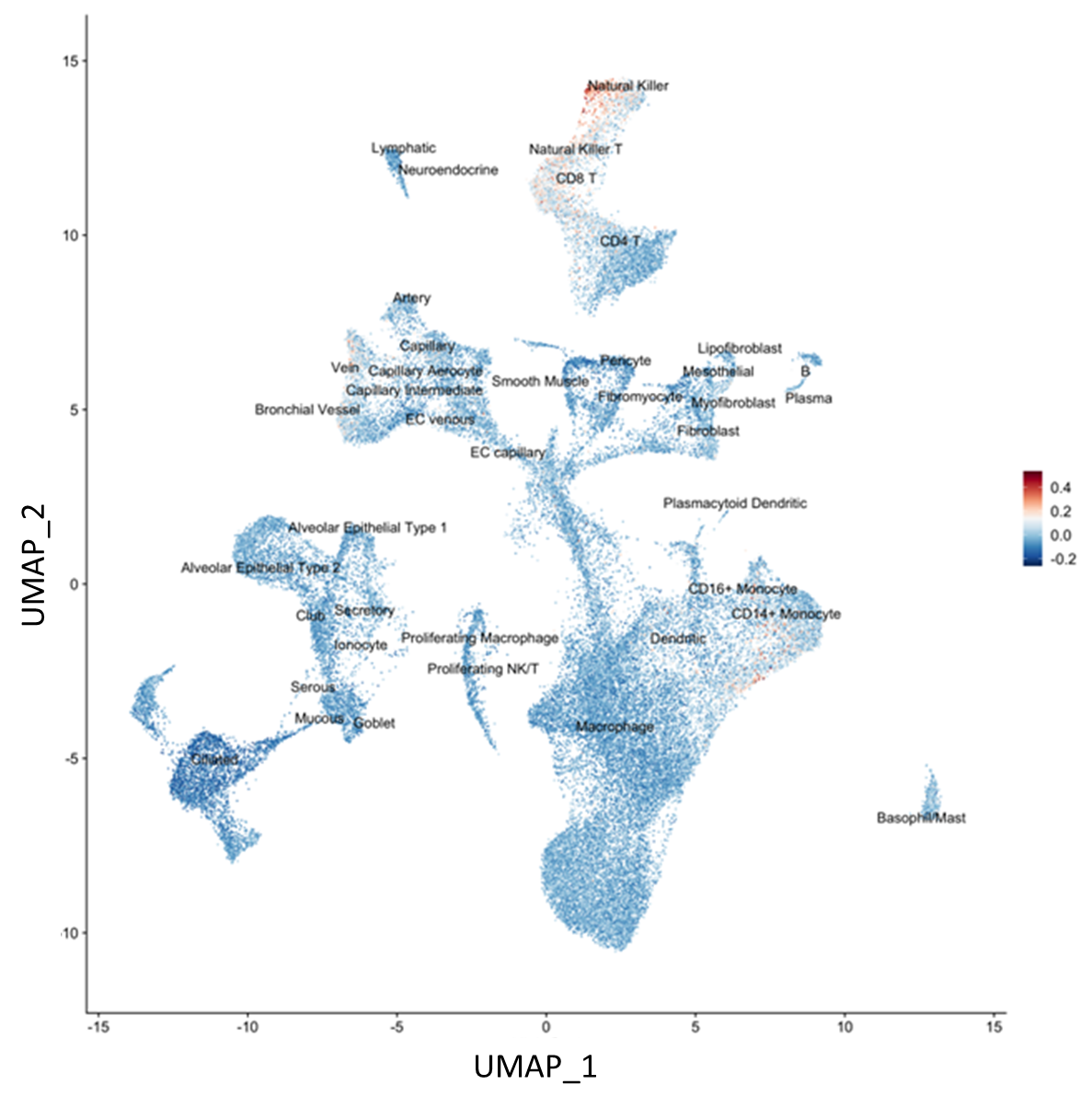

**Figure S1.** Uniform manifold approximation and projection (UMAP) of single cell RNA sequencing from lungs of patients with SSc-ILD (derived from GEO dataset GSE128169) demonstrated enrichment of genes associated with cGAS expression across multiple cell populations.
