## Supplemental Figure 2 for "cGAS Expression is Enhanced in Systemic Sclerosis Associated Interstitial Lung Disease and Stimulates Inflammatory Myofibroblast Activation"

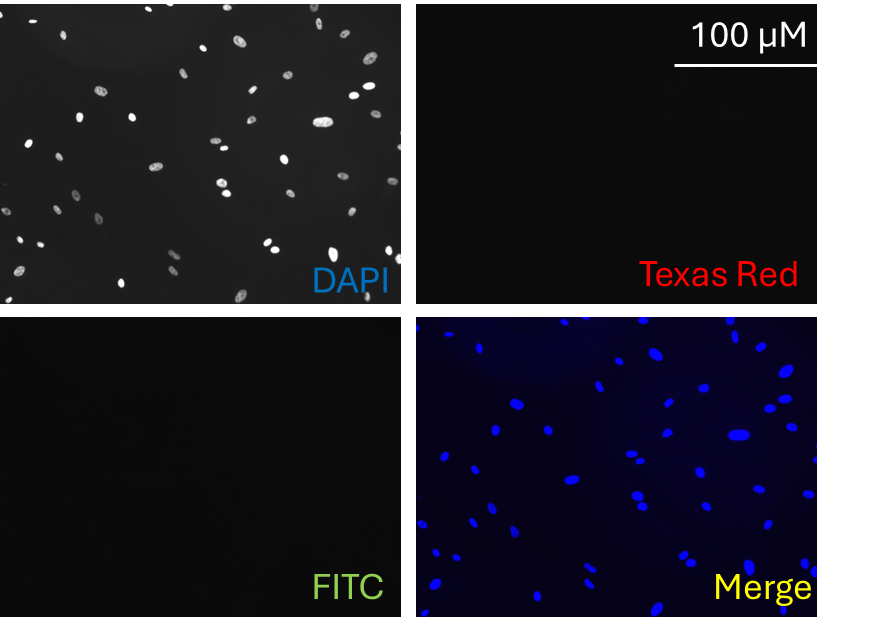

**Figure S2. Negative control for immunofluorescence staining.** Sections stained with goat anti-mouse IgG (Texas Red, Alexa Fluor® 555 conjugate) and donkey anti-rabbit IgG (FITC, Alexa Flour® 488 conjugate) secondary antibodies in the absence of primary antibody show minimal signal in the red, green, and merged (yellow) channels. Slides are counterstained with DAPI (blue). Scale bar = 100 microns.
