## Supplemental Figure 3 for "cGAS Expression is Enhanced in Systemic Sclerosis Associated Interstitial Lung Disease and Stimulates Inflammatory Myofibroblast Activation"

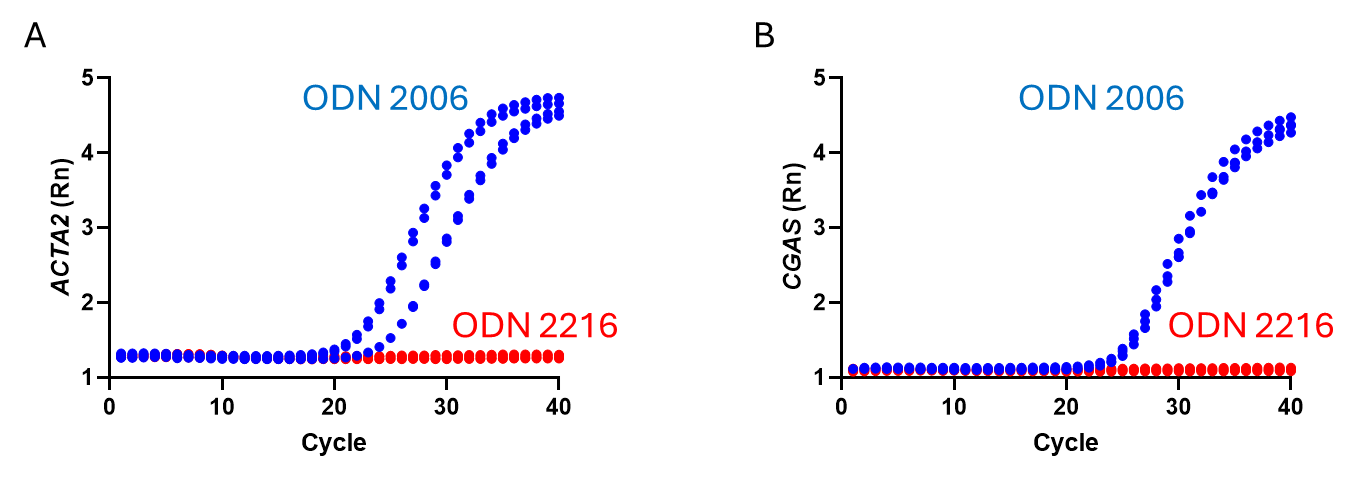

**Figure S3. Co-stimulation of NHLFs with TGFβ1 and CpG ODN2006 induces *ACTA2* and *CGAS* transcription.** qRT-PCR amplification curves of TGFβ1-treated NHLFs subjected to exogenous CpG ODN2006 (blue) or ODN2216 (red). Administration of CpG ODN2006 (but not OD2216) resulted in the transcription of **(A)** *ACTA2* and **(B)** *CGAS*.
