## Supplemental Figure 4 for "cGAS Expression is Enhanced in Systemic Sclerosis Associated Interstitial Lung Disease and Stimulates Inflammatory Myofibroblast Activation"

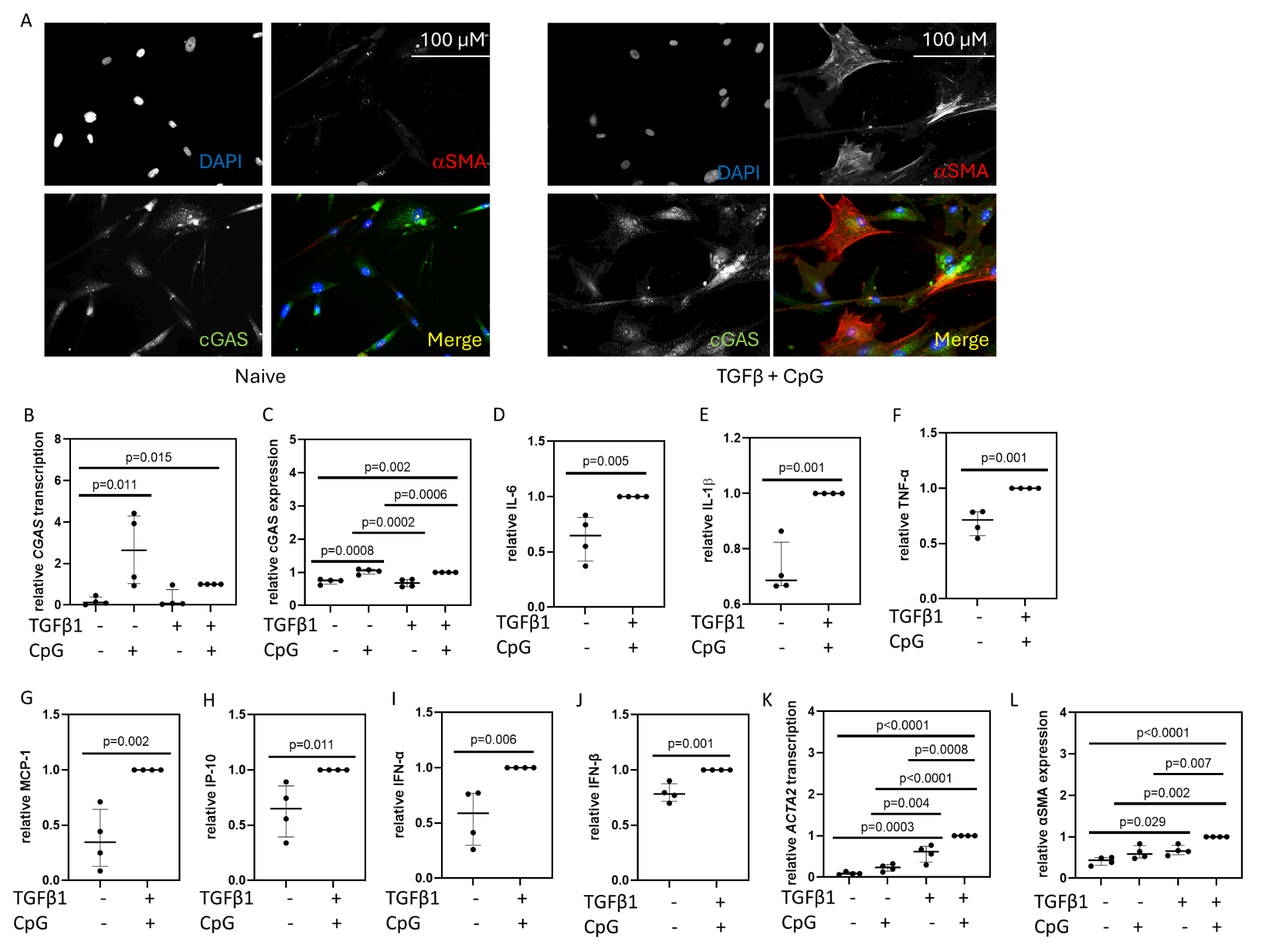

**Figure S4. Co-stimulation of NHLFs with TGFβ1 and CpG induces cGAS activation. (A)** IF of naïve (left) and TGFβ1 and CpG ODN2006 co-stimulated (right) NHLFs showed the cytoplasmic aggregation of cGAS in stimulated cells. Scale bar = 100 microns. These stimulated cells displayed increased mRNA transcription and protein expression of **(B,C)** cGAS, high levels of **(D)** IL-6, **(E)** IL-1β, **(F)** TNF-α, **(G)** MCP-1, **(H)** IP-10, **(I)** IFN-α, and **(J)** IFN-β expression, and elevated transcription and expression of **(K,L)** αSMA. Data was normalized to the TGFβ1 + CpG condition.
