## Supplemental Figure 5 for "cGAS Expression is Enhanced in Systemic Sclerosis Associated Interstitial Lung Disease and Stimulates Inflammatory Myofibroblast Activation"

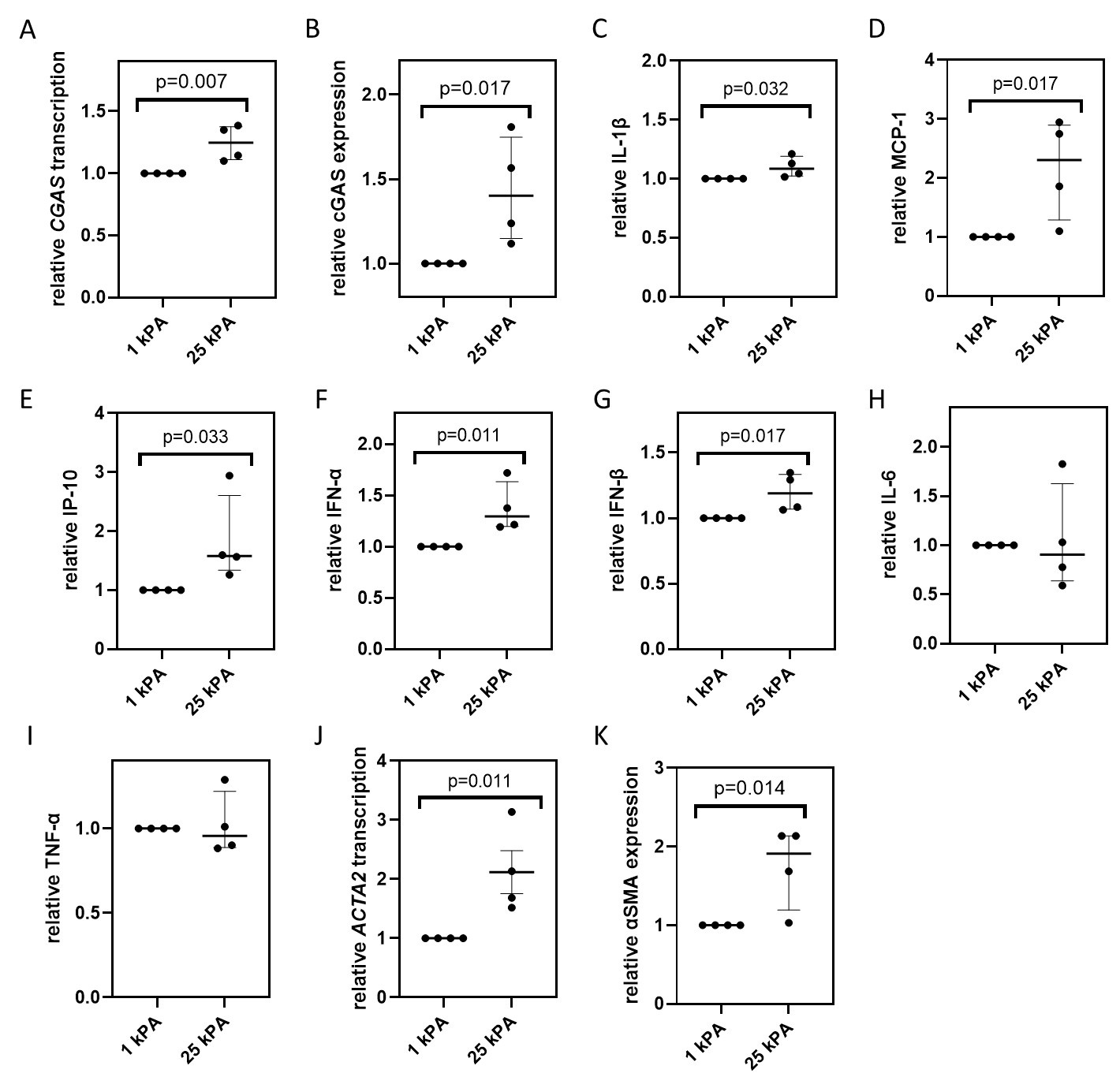

**Figure S5. Stiff substrates induce expression of cGAS and inflammatory fibrotic responses in NHLFs.** Relative to NHLFs cultured on hydrogels that approximate the normal lung (1 kPA), cells cultured on hydrogels that simulate the fibrotic lung (25 kPA) exhibited increased transcription and protein expression of **(A,B)** cGAS that was associated with elevated levels of **(C)** IL-1β, **(D)** MCP-1, **(E)** IP-10, **(F)** IFN-α, and **(G)** IFN-β; **(H)** IL-6 and **(I)** TNF-α expression were unchanged between conditions. **(J,K)** αSMA mRNA and protein levels appropriately rose in response to the 25 kPA condition. Data were normalized to the 1 kPA condition.
