## Supplemental Figure 6 for "cGAS Expression is Enhanced in Systemic Sclerosis Associated Interstitial Lung Disease and Stimulates Inflammatory Myofibroblast Activation"

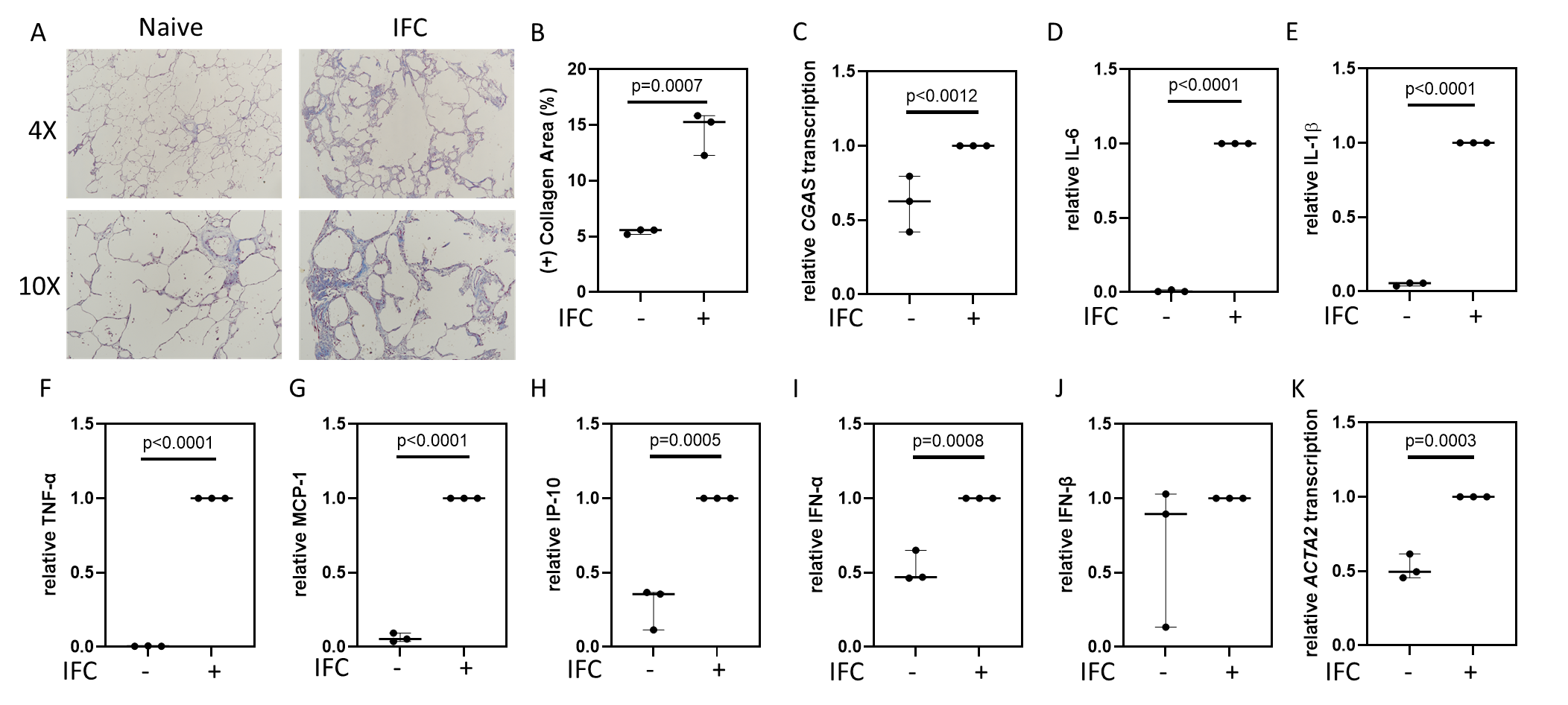

**Figure 6S. An inflammatory fibrotic cocktail induces fibrosis-like changes in a human PCLS model. (A)** Representative images of Masson’s Trichrome staining of human PCLS sections from those subjected to normal media or an inflammatory fibrotic cocktail (IFC). **(B)** Collagen quantification via ImageJ software revealed increased collagen content following administration of this IFC. Additionally, PCLS subjected to this IFC exhibited elevated transcription of **(C)** *CGAS* and protein levels of **(D)** IL-6, **(E)** IL-1β, **(F)** TNF-α, **(G)** MCP-1, **(H)** IP-10, and **(I)** IFN-α. This IFC did not influence expression of **(J)** IFN-β. As expected, this IFC induced transcription of **(K)** *ACTA2* Data were normalized to the IFC condition.
