## Supplemental Figure 7 for "cGAS Expression is Enhanced in Systemic Sclerosis Associated Interstitial Lung Disease and Stimulates Inflammatory Myofibroblast Activation"

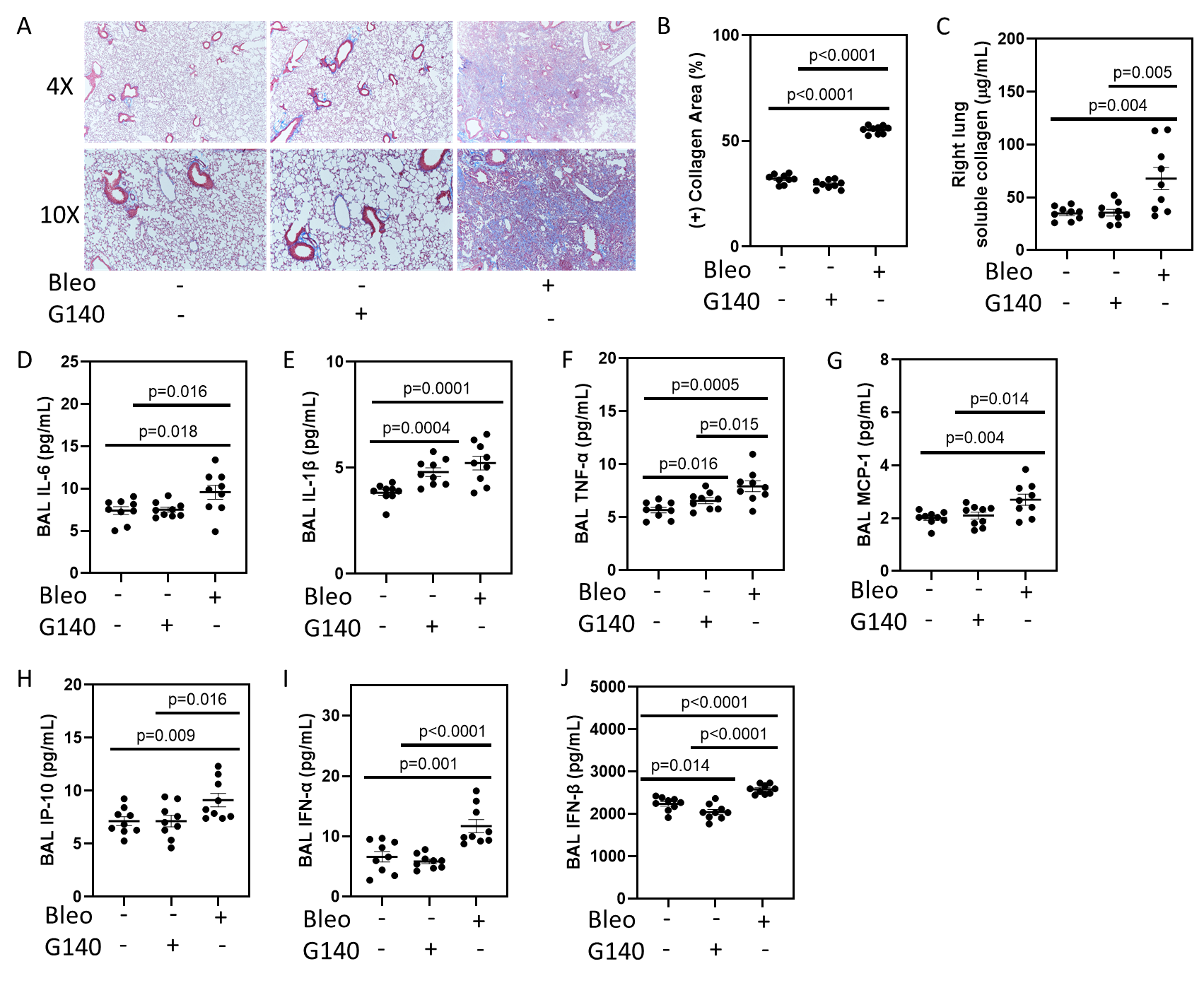

**Figure S7. Bleomycin exposed mice display increased lung collagen content and BAL cytokine, chemokine, and type 1 interferon concentrations.** C57Bl/6 mice received intraperitoneal injections of G140 or normal saline or a single dose of inhaled bleomycin or normal saline. **(A)** Representative images of Masson’s Trichrome staining of mouse lung sections. Collagen quantification via **(B)** ImageJ software and **(C)** Sircol assay revealed increased collagen content in bleomycin challenged mice as compared to naïve or G140 treated mice. Similarly elevated BAL levels of **(D)** IL-6, **(E)** IL-1β, **(F)** TNF-α, **(G)** MCP-1, **(H)** IP-10, **(I)** IFN-α, and **(J)** IFN-β were observed in bleomycin exposed mice.
