## Supplemental Methods for "cGAS Expression is Enhanced in Systemic Sclerosis Associated Interstitial Lung Disease and Stimulates Inflammatory Myofibroblast Activation"

**SUPPLEMENTARY METHODS**

**Single Cell RNA Sequencing (scRNA seq) Analysis**

Analyses of scRNA seq data from GSE128169 were performed with Seurat version 5.0.318. A Seurat object was created for this dataset and data was normalized. Mitochondrial and cell cycle genes were identified and used for adjustments in this dataset. RNAseq annotation was performed with Azimuth [1]. Principal components analysis was performed, and 70 significant principal components were chosen using Jack Straw for downstream cell clustering and visualization using the uniform manifold approximation and projection (UMAP) technique. To identify cell types expressing genes associated with cGAS, the AddModuleScore, a Seurat function that calculates the gene expression level of a cluster of genes on a single cell level and then subtracts the aggregated expression of a randomly assigned control set, was used. These cGAS genes were visualized using the UMAP technique as well as Violin plots with bootstrap for nonparametric confidence intervals.

**Human Subjects**

Human studies were conducted with informed consent and with protocols approved by the Institutional Review Board at the Yale School of Medicine (HIC2000024862). Diagnosis of scleroderma (SSc) and scleroderma-associated interstitial lung disease (SSc-ILD) were based on consensus guidelines following multidisciplinary discussion [2, 3]. Healthy control (n=12) and SSc patients lacking (n-8) and having (n=15) ILD were recruited from the Yale ILD Center of Excellence and Yale Scleroderma Program from 2010-2022 with demographic and clinical data obtained upon enrollment. These data included age, gender, race, smoking history, SSc subtype (diffuse or limited), disease duration, modified Rodnan skin score (MRSS), presence or absence of autoantibodies (anti-Scl-70, centromere, or RNA Polymerase III), and pulmonary function testing for percent predicted forced vital capacity (FVC%) and percent predicted diffusion capacity for carbon monoxide (DLCO%). Bronchoalveolar lavage (BAL) was collected with the following methods. Serial lavages were performed by instillation of 30-50 ml of sterile 0.9% normal saline into right middle lobe or lingula of the lung for a maximum of 150 ml. The resultant BAL samples were subject to two centrifugations at 1000 RPM of 30 minutes to ensure separation of the cellular pellet. The cell-free supernatant was aliquoted and stored at -80C until the time of analysis.

**Cell Culture**

Normal human lung fibroblasts (NHLFs) were cultured in a humidified 5% CO2 incubator at 37°C in medium made up of Dulbecco’s Modified Eagle’s Medium (DMEM, ThermoFisher) supplemented with 10% fetal bovine serum (FBS, ThermoFisher) and 1% penicillin/streptavidin (Pen-Strep, ThermoFisher). After 3-5 passages, cells were seeded onto either standard 6-well plates (Corning) for biochemical stimulation or tunable collagen-based hydrogels (Matrigen) for biomechanical studies. *Biochemical stimulation:* Cells were seeded onto 6-well plates, approximately 100,000 cells/well. After a 24 hour period of serum starvation, cells were stimulated with 5 ng/ml of recombinant human TGFβ1 (R&D Systems) and 50 µM of CpG DNA (Invivogen) for 48 hours. TGFβ1-and CpG-treated cells were then treated to the presence or absence of 10 µM of G140 (Invivogen) for 24 hours. *Biomechanical studies:* NHLFs were seeded onto hydrogels of 1 kPa and 25 kPa stiffness to simulate the rigidity of the normal and fibrotic lung, respectively. These cells were cultured for seven days and then subjected to the absence or presence of 10 µM of G140 for 24 hours. Fibroblasts harvested from SSc-ILD lungs were cultured media containing DMEM, 10% FBS, and 1% Pen-Strep, passaged 3-5 times, seeded onto standard 6-well plates, and treated in the absence or presence of G140 for 24 hours. For all experiments, cells were harvested for immunofluorescence, RNA, and protein studies.

**Mouse Studies**

Mouse studies were conducted in accordance with procedures approved by the Institutional Animal Care and Use Committee at Yale School of Medicine. C57BI/6 mice, aged 9-11 weeks, received a single dose of inhaled bleomycin (0.8 units/kg, McKesson) or normal saline. On days 3, 7, and 10 post-bleomycin inhalation, mice were treated with vehicle control or G140 (30 mg/kg) via intraperitoneal injection. Mice were euthanized on day 14 for collection of lungs and BAL; after terminal anesthetization, BAL was performed, followed by median sternotomy and right heart perfusion. Lungs were harvested *en bloc* and processed for the studies described below.

**Precision cut lung slices (PCLS)**

Normal human PCLS were obtained from healthy donors from the Institute for In Vitro Sciences. Upon being rapidly thawed in 70% ethanol, PCLS were cultured according to the manufacturer protocol in acclimation media made up of DMEM: F12 (Lonza) with 0.2% Primocen® (Invivogen), 1% insulin-transferrin-selenium (ThermoFisher), 1% Antibiotic Antimycotic solution (Millipore Sigma), 2µM hydrocortisone (Millipore Sigma), and 2-phospho-L-ascorbic acid trisodium salt (Millipore Sigma). After three days in the acclimation media, PCLS were cultured in culture media comprising of DMEM: F12 with 0.2% Primocen® and 1% insulin-transferrin-selenium for another three days. Next, PCLS were treated in the absence or presence of an inflammatory fibrotic cocktail (IFC) for five days, which consisted of 5 ng/ml of TGFβ1, 5 μM of platelet-derived growth factor-AB (ThermoFisher), 10 ng/ml of tumor necrosis factor alpha (TNFα, R&D Systems), 5 μM of lysophosphatidic acid (Cayman Chemical), and 50 µmol of CpG. PCLS subjected to this IFC were then treated in the absence or presence of 10 µM of G140 for 24 hours. PCLS were harvested for histology, RNA, and protein studies.

**RNA Isolation and RT-qPCR**

Total cellular RNA was extracted from fibroblasts and PCLS harvested in Qiazol (Qiagen). RNA Isolation was performed with the miRNeasy Mini-Kit (Qiagen) as per the manufacturer’s protocol. One-step RT-qPCR utilizing primers for alpha smooth muscle actin (αSMA, ThermoFisher), cGAS (ThermoFisher), and GAPDH (ThermoFisher) was then conducted on the ViiA 7 Real-Time PCR System (ThermoFisher) with the Applied Biosystems TaqMan RNA-to-CT 1-Step Kit (ThermoFisher). Relative expression of αSMA and cGAS was calculated using the 2-delta Ct method as previously described [4, 5].

**Cell Lysis**

Cells and PCLS were washed with PBS and lysed in a solution containing Pierce RIPA buffer (ThermoFisher) and a Halt™ Protease Inhibitor Cocktail (ThermoFisher).

**Quantification of Cytokines**

Human BAL, cell lysates, PCLS, or mouse BAL was subject to multiplex quantification of the following cytokines: monocyte chemoattractant protein-1 (MCP-1), tumor necrosis factor alpha (TNFα), interferon-gamma inducible protein-10 (IP-10), interleukin-6 (IL-6), interleukin 1-beta (IL-1β), interferon alpha (IFNα), and interferon beta (IFNβ) using the U-PLEX Cytokine Panel Human Kit (Mesoscale Discovery) or U-Plex Cytokine Panel Mouse Kit (Mesoscale Discovery) per the manufacturer’s protocol. Briefly, samples were diluted in a 1:2 ratio and plated in duplicate. Cytokine concentrations were quantified as pg/mL using the Meso Scale Discovery QuickPlex SQ 120 Model 1300 with the Methodical Mind software (Meso Scale Discovery).

**αSMA ELISA**

Cell lysates were assayed for αSMA via commercially available ELISA (Abcam) as per the manufacturer’s protocol. Briefly, samples were diluted in 1:6 ratio and plated in duplicated. Concentrations of αSMA were quantified as pg/mL at an absorbance 450 nm using the Gen5™ Microplate Data Collection and Analysis Software (BioTek Instruments).

**cGAS ELISA**

Cell lysates were assayed for cGAS via commercially available ELISA (Cell Signaling Technology) as per the manufacturer’s protocol. Briefly, samples were diluted in 1:5 ratio and plated in duplicated. Concentrations of αSMA were quantified as pg/mL at an absorbance 450 nm using the Gen5™ Microplate Data Collection and Analysis Software (BioTek Instruments).

**MTT Assay**

NHLFs were detached with 0.05% trypsin/EDTA, counted, and approximately 10,000 cells were seeded onto 96-well cell culture plates. Cells were labeled with 12 nM MTT (ThermoFisher), lysed with DMSO, and analyzed at an absorbance 540 nm using the Gen5™ Microplate Data Collection and Analysis Software (BioTek Instruments).

**Immunohistochemistry (IHC)**

Formalin fixed, paraffin embedded slides of normal or SSc lung sections from human donors were a kind gift from Dr. Carol Feghali-Bostwick. Slides were deparaffinized in xylene and rehydrated to distilled water in graded ethanol. An antibody for cGAS (Invitrogen) diluted in a 1:100 ratio was applied for 60 minutes. The slides were washed in Tris buffer and horseradish peroxidase (HRP)-conjugated anti-rabbit (BioCare Medical) were applied for 30 minutes. Slides were washed with Tris buffer, developed with DAB, counterstained with hematoxylin, and cover-slipped with a resinous mounting media. Images were acquired with a Nikon D5-Ri2 microscope. Quantification of cGAS was performed by manually counting total nuclei and nuclei from cGAS-expressing cells from normal (n=4) and SSc-ILD (n=4) lung slides. The percentage positive cGAS cells per high-power field were reported.

**Immunofluorescence (IF)**

Immunofluorescence detection of αSMA and cGAS in SSc fibroblasts were performed using the following primary antibodies: mouse anti-αSMA (1:250, Abcam) and rabbit monoclonal cGAS (1:100, Invitrogen). This was followed by secondary antibody detection and DAPI counterstaining. Negative controls included slides that were processed and detected in the absence of primary antibody. Images were acquired with a Zeiss confocal microscope.

**Lung Histology and Collagen Quantification**

Formalin fixed, paraffin embedded sections from PCLS or whole left lungs from mice were stained with Masson’s trichrome to visualize collagen deposition as previously reported [6]. All sections were analyzed using a Nikon D5-Ri2 microscope. *Quantification of collagen* was done via ImageJ software using color deconvolution 2 plugin. Briefly, microscopic scanning of the slides was conducted in a bright field with a Nikon D5-Ri2 inverted microscope at 20X magnification. Four random, non-overlapping fields of view were used from each section for collagen quantification, which was calculated as the trichrome positive area divided by the total area. *Mouse lungs* were snap frozen in liquid nitrogen and stored at −80°C until quantification of soluble collagen using the Sircol Collagen Assay kit (Biocolor) as previously reported [7].

**Bulk RNA Sequencing**

*RNA Seq Library Prep:* Using the Kapa RNA HyperPrep Kit with RiboErase (KR1351), rRNA was depleted starting from 25-1000ng of total RNA by hybridization of rRNA to complementary DNA oligonucleotides, followed by treatment with RNase H and DNase to remove rRNA duplexed to DNA. Samples were fragmented using heat and magnesium. 1st strand synthesis was performed using random priming. 2nd strand synthesis incorporated dUTPs into the 2nd strand cDNA. Adapters were ligated and the library was amplified. Strands marked with dUTPs were not amplified, allowing for strand-specific sequencing. Indexed libraries that met appropriate cut-offs for both quantity and quality were quantified by qRT-PCR using a commercially available kit (KAPA Biosystems) and insert size distribution was determined with the LabChip GX or Agilent Bioanalyzer. Samples with a yield of ≥0.5 ng/µl were used for sequencing. *Flow Cell Preparation and Sequencing:* Sample concentrations were normalized to 1.2 nM and loaded onto an Illumina NovaSeq flow cell at a concentration that yielded 80 million passing filter clusters per sample. Samples were sequenced using 100bp paired-end sequencing on an Illumina NovaSeq according to Illumina protocols. The 10bp unique dual index was read during additional sequencing reads that automatically followed the completion of read 1. Data generated during sequencing runs were simultaneously transferred to the Yale Center for Genome Analysis (YCGA) high-performance computing cluster. A positive control (prepared bacteriophage Phi X library) provided by Illumina was spiked into every lane at a concentration of 0.3% to monitor sequencing quality in real time. *Data Analysis and Storage:* Signal intensities were converted to individual base calls during a run using the system's Real Time Analysis (RTA) software. Base calls were transferred from the machine's dedicated personal computer to the Yale High Performance Computing cluster via a 1 Gigabit network mount for downstream analysis. Primary analysis - sample de-multiplexing and alignment to the human genome - was performed using Illumina's CASAVA 1.8.2 software suite. Adapter sequences were removed using Cutadapt, and reads were aligned using STAR to human genome.

**Statistical Analysis**

Data distribution was assessed using Shapiro-Wilk test, and categorical data was analyzed with Fisher’s exact test using GraphPad Prism 9.4.1. Multiple hypothesis testing was conducted with one-way ANOVA with Bonferroni correction with GraphPad Prism. Pairwise comparisons were conducted with an unpaired t-test or Mann Whitney test based on data distribution with GraphPad Prism. p-values < 0.05 were considered significant.
